# Third places visits and well-being: insights from longitudinal passive sensing data

**DOI:** 10.64898/2026.03.12.26348188

**Authors:** Yu Fang, Kevin Saulnier, Jennifer Cleary, Zhenke Wu, Amy Bohnert, Srijan Sen

## Abstract

Spending time in locations outside the home and workplace (termed “third places”), has been linked to better mental health. However, studies to date have typically been cross-sectional, based on self-reported location data and employed small sample sizes, limiting their ability to assess the presence and nature of the association between third places and mental health. To overcome these limitations, we collected 18,795 person-days of objective SensorKit location data passively from a national cohort of 410 first-year medical residents across the United States, to assess visits to third places and their associations with mood and depression over the course of one year. On average, participants visited 3.3 unique locations per day (SD=1.7) and spent 17.9% of their time at third places (SD = 26.5%). Within individuals, both a higher percentage of time spent at third places (B=0.013 [per 10% increase], p<0.001) and a greater number of unique locations visited (B=0.032, p<0.001) were associated with better mood later that same day, independent of the time spent at work. These associations were partially mediated by step counts and outdoor light exposure jointly (19.2% and 27.6%). Reverse-direction associations were observed, with better mood on one day predicting both more time spent at third places (B=0.052, *p*<0.001) and more unique locations visited (B=0.032, *p*<0.001) the following day. Between subjects, depressed subjects spent less percentage of time at third places (12.3% vs. 21.2%, t=-3.7, *p*<0.001) and visited fewer unique places per day (2.9 vs. 3.4, t = –3.8, *p*<0.001) compared to non-depressed subjects. These findings demonstrate the relationship between visiting third places and well-being, and suggest that interventions and policies aimed at encouraging third places visits have the potential to improve mental health.

## Introduction

The term “third places”^1^ describes locations distinct from home (the “first place”) and workplace (the “second place”), such as cafes, parks, community centers, and libraries^2^. Scholars have theorized that third places support social connection^3–7^ and community attachment^7–9^, a stronger sense of belonging, and potentially a lower risk of depression and anxiety^10,11^. In addition, third places like parks^12–18^ and gyms provides opportunities for exposure to outdoor environments and sunlight^19^ and physical activity^20,21^, experiences that are associated with psychological well-being^22–25^. Taken together, these findings suggest that third places are a promising target for well-being interventions, and could potentially act through social, environmental, and activity-driven mechanisms.

While the theoretical basis supporting the potential importance of third places in mental health and wellbeing is compelling and well-developed, the accompanying supporting empirical evidence remains comparatively limited. Existing empirical research has been constrained by several limitations. First, most studies have relied on subjective, self-reported location data such as questionnaires^3–5,8,15^ or photographs^25^, which are susceptible to recall and reporting biases. This dependence on self-report methods has also led to a narrow focus on specific types of third places, such as coffee shops^4^, libraries^9^, and museums^26^, rather than capturing the full spectrum of third places. Though recent advances in passive sensing technologies promise to overcome these limitations, only a handful of studies have incorporated such methods^13,25,27^, and these remain limited by small sample sizes (fewer than 100 participants)^25,27^, limited settings (e.g., only assessing homestays)^27^ and brief data collection periods spanning just a few days^13^. Furthermore, prior studies investigating the relationship between third places and mental health predominantly utilized either cross-sectional^3–5,7,8,13–15,17,24,25^ or simple pre-post designs^27^. These methodological and study design limitations substantially restrict the ability to assess within-person changes in third places visits and their dynamic relationship with mental health outcomes over time, which is essential to inform individual-level intervention strategies; and underscore the need for large-scale, longitudinal studies utilizing passive, objective, and comprehensive measures of third places engagement together with repeated assessment of mental health outcomes.

In the present study, we hypothesized that more third places visits would show a bidirectional association with better mood within individuals, and that this relationship would be partially mediated by factors such as physical activity and light exposure. Additionally, at the between-person level, we hypothesized that there would be fewer third places visits among depressed individuals than non-depressed individuals. To test these hypotheses, We collected objectively-measured third places visits, physical activity, and light exposure, alongside daily self-reported mood, and quarterly PHQ-9 depressive symptoms assessments across a large sample of subjects across a full year. Third places visits data was passively recorded using the “Visits” sensor^28^ from SensorKit (Apple Inc.)^29,30^, a privacy-preserving tool for longitudinal assessment of location behaviors at scale. With these methodological strengths, we sought to identify whether third places visits are associated with mental health, both between and within individuals, and identify possible mechanisms linking third places to mental health.

## Methods

### Study Design

The Intern Health Study is an annual cohort study that follows first-year physician residents (interns) across the United States. Interns starting residency in 2023 were invited to participate via email 2 to 3 months before the start of internship (in July of 2023). Upon enrollment, participants were provided with their choice of wearable devices including Fitbit Charge 4, Inspire 2 (Google Fitbit, San Francisco, CA), Apple Watch (Apple Inc., Cupertino, CA), or $50 if they already owned an eligible wearable (Fitbit, Apple watch or Garmin watch [Garmin Ltd., Olathe, KS]). Interns received up to $80 additional compensation throughout the year for completing quarterly surveys. Participants using iPhones for participation were asked to enroll in the SensorKit arm of the study through additional consenting processes^31^. No additional incentives were provided for opting into the collection of SensorKit data. The institutional review board at the University of Michigan approved the study (HUM00033029).

### Data Collection

#### Demographics and depressive symptoms

Upon enrollment, participants completed a baseline survey via the study app (MyDataHelps, CareEvolution, Ann Arbor, MI) assessing demographic information, including age, sex, medical specialty, relationship and parental status. The baseline survey, as well as quarterly surveys administered across the internship year, also assessed depressive symptoms via the 9-item Patient Health Questionnaire (PHQ-9)^32,33^. The PHQ-9 assesses whether during the previous 2 weeks, a set of 9 depressive symptoms had bothered subjects “not at all”, “several days”, “more than half the days”, or “nearly every day”. The diagnostic validity of the PHQ-9 is similar to that of clinician-administered assessments^33,34^.

#### Daily mood

Before and throughout the intern year, participants received push notification from the study app at a user-specified time between 5:00 pm and 10:00 pm (default 8:00 pm) each day, prompting them to answer the question: “On a scale of 1 (lowest) to 10 (highest) how was your mood today?”. Participants could also access the app independently to complete the mood survey at any time, multiple times per day.

#### Location data

We utilized SensorKit to identify visited locations. SensorKit is a technological framework that enables researchers to collect relevant sensor data from consented participants via a study app that reads the data from iPhones and Apple Watches. SensorKit Visits sensor provides the date and time of arrival and departure of each visited location, with the times grouped to the nearest 15-minute increments (i.e., daily at 00:00, 00:15, …, 23:30, 23:45, 24:00) in order to preserve subject privacy. Each location is categorized as ‘home’, ‘work’, ‘school’, ‘gym’ and ‘other’ through a hybrid process combining automated system detection with manual labeling by participants in their iOS Apps. Each unique geographic location is assigned an anonymized, 36-character string identifier. For privacy protection, specific GPS coordinates, location names, and addresses of the locations (including home and work) were not collected (**Table S1**).

#### Additional variables

Ambient light data and daily step counts were also collected for mediation analyses outlined in the Statistical Analyses sections. Ambient light data was collected through SensorKit Ambient Light sensor^35^, which records illuminance (in the unit of LUX) in subjects’ environment. Accelerometer-based daily step counts were collected through wearable wrist-band devices (Fitbit, Apple Watch, or Garmin) from subjects.

### Statistical Analyses

All statistical analyses were conducted with the use of R version 4.3.2 (The R Foundation, Vienna, AUT)^36^.

#### Data preprocessing

We included the location data across the full intern year (from July 2023 to June 2024) in analyses. Days with less than 50% coverage (i.e., fewer than 12 hours of location data) were excluded. From the original location data, we derived three daily metrics that reflect participants’ place visiting behavior: 1) the percentage of time spent at third places (places outside the home and workplace), calculated by the proportion of time recorded at locations with the type of ‘other’ relative to the total time with recorded location data; 2) the number of unique visited places (including all types), calculated by the number of unique anonymized identifiers; and 3) the percentage of time spent at workplace (locations labeled as ‘work’) each day.

The ambient light illuminance data were sampled at 10-minute intervals. To assess daily outdoor light exposure, we calculated the percentage of time with light exposure illuminance levels >= 1000 lux (the mean illuminance of an overcast day^37,38^) of each day.

To enhance interpretability of effect sizes, variables expressed as percentages (the percentage of time spent at third places, the percentage of time spent at the workplace, and the percentage of time with light exposure at illuminance levels ≥1000 lux) were divided by 10 in the statistical analyses. As a result, all reported effect sizes reflect the estimated change associated with a 10% increase in these variables.

Participants with at least three days of complete data, including mood ratings, location visits, outdoor light exposure, step counts, and previous day’s mood, through the intern year, were included in the final analyses.

#### Third places visits and daily mood

We utilized linear mixed modeling^39^ to assess the relationships between third places visits and daily mood, with a random intercept to account for repeated measures of individuals. We first investigated whether the percentage of time spent at third places and the number of unique places visited were associated with mood later that day. To further disaggregate between-subject and within-subject effects of both the percentage of time spent at third places and the number of unique places, we calculated each participant’s average values as the between-person variables and then derived the within-person variables by subtracting the individual average values from each daily value.

To determine the demographic covariates to include in final models, we assessed the bivariate associations between demographic measures (age, sex, medical specialty [surgical or non-surgical], relationship status [single or non-single] and parental status [having kids or not]) with the third places visits, and retained those with significant associations. We also included weekend/weekday, average daily mood before internship started and previous day’s mood as covariates.

Based on prior findings^40,41^, longer work hours on a given day can be associated with lower mood, along with less time available for visiting third places, and thus was a potential confounder. To assess whether the observed effects of visiting third places were attributable to time not working, we repeated the models, adding the percentage of time spent at the workplace as an additional covariate.

Days on which participants spent none or all of their time at third places (e.g., sick day exclusively spent at home or a vacation day exclusively spent at third places) may represent qualitatively different circumstances compared to days with some time spent there. To assess whether results were driven by these “none” or “all” days, we created a categorical variable based on the percentage of time spent at third places with levels of “No third places” (0% time spent at third places), “All third places” (100% time spent at third places), and “Some third places” (0% < time spent at third places < 100%, used as the reference level). Then we repeated the analysis replacing the continuous variable of the percentage of time spent at third places with this categorical variable. We also repeated the analysis with the continuous variable but excluding days of “No third places” or “All third places”.

Next, we examined whether higher mood on one day was associated with measures of third places visits the following day (i.e., reverse relationship). We constructed two models, one with the percentage of time spent at third places as the outcome and one with the number of unique places visited as the outcome. Both models used daily mood from the previous day as the explanatory variable, adjusted for sex, specialty, weekend/weekday, and included a random intercept for each participant. Additionally, we repeated the analysis for the percentage of time spent at third places, excluding days of “No third places” or “All third places”. To address skewness of the percentage of time spent at third places (Pearson’s coefficient of skewness = 1.84) and the number of unique places visited (Pearson’s coefficient of skewness = 1.65), we applied a rank-based inverse normal transformation to these two variables.

#### Mediation effects of step counts and outdoor light exposure on the relationship between third places visits and mood

We assessed the independent mediation effects of physical activity and outdoor light exposure on the relationship between third places and mood, and subsequently assessed their combined mediation effect using structural equation modeling^42^. Daily step counts and daily percentage of time with illuminance >= 1000 lux were used as measures of physical activity and outdoor light exposure, respectively. Baseline average daily mood, previous day mood, sex, medical specialty, weekend/weekday, and percentage of time spent at workplace were included as covariates.

#### Third places visits comparison between subjects who met criteria for depression and those who did not

We investigated whether individuals whose average PHQ-9 scores over internship year was greater than or equal to 10, a threshold which has moderate sensitivity (88%) and specificity (85%) for a diagnosis of depression^43^, differed in third places visits compared to subjects who did not meet this threshold. We conducted independent-sample t-tests of the quarterly means of 2-week average percentage of time spent at third places and number of unique places visited between the depressed and non-depressed groups. To account for potential confounding by baseline PHQ-9 score, sex, and medical specialty, we also estimated and compared the marginal group means of the third places outcomes for the two groups by adjusting for these covariates.

#### Sensitivity analysis

Third places visits during night time may indicate overnight stays at a partner or a friend’s residence, which may not align with the intended definition of third places visits in this study. Therefore, we conducted a sensitivity analysis where records from midnight to 6 a.m. were excluded. This exclusion left a maximum of 18 hours of records per day, and only days with >=50% coverage (9 hours) were included.

## Results

### Sample Characteristics

A total of 4151 interns starting residency in July 2023 were invited to participate in the 2023 Intern Health Study and 1437 interns enrolled in the study. Among them, 1164 participants were iPhone users (81%). After further excluding participants who opted out of SensorKit Visits data collection, those who did not have both home and work locations labeled, and those without sufficient data, 410 participants were included in the final analyses (see **Figure 1** for the details of study flow). Of included participants, 223 (54.3%) were female, their mean age was 27.7 years (SD = 2.7), and 74 (18.0%) were in surgical specialties.

**Figure 1.**
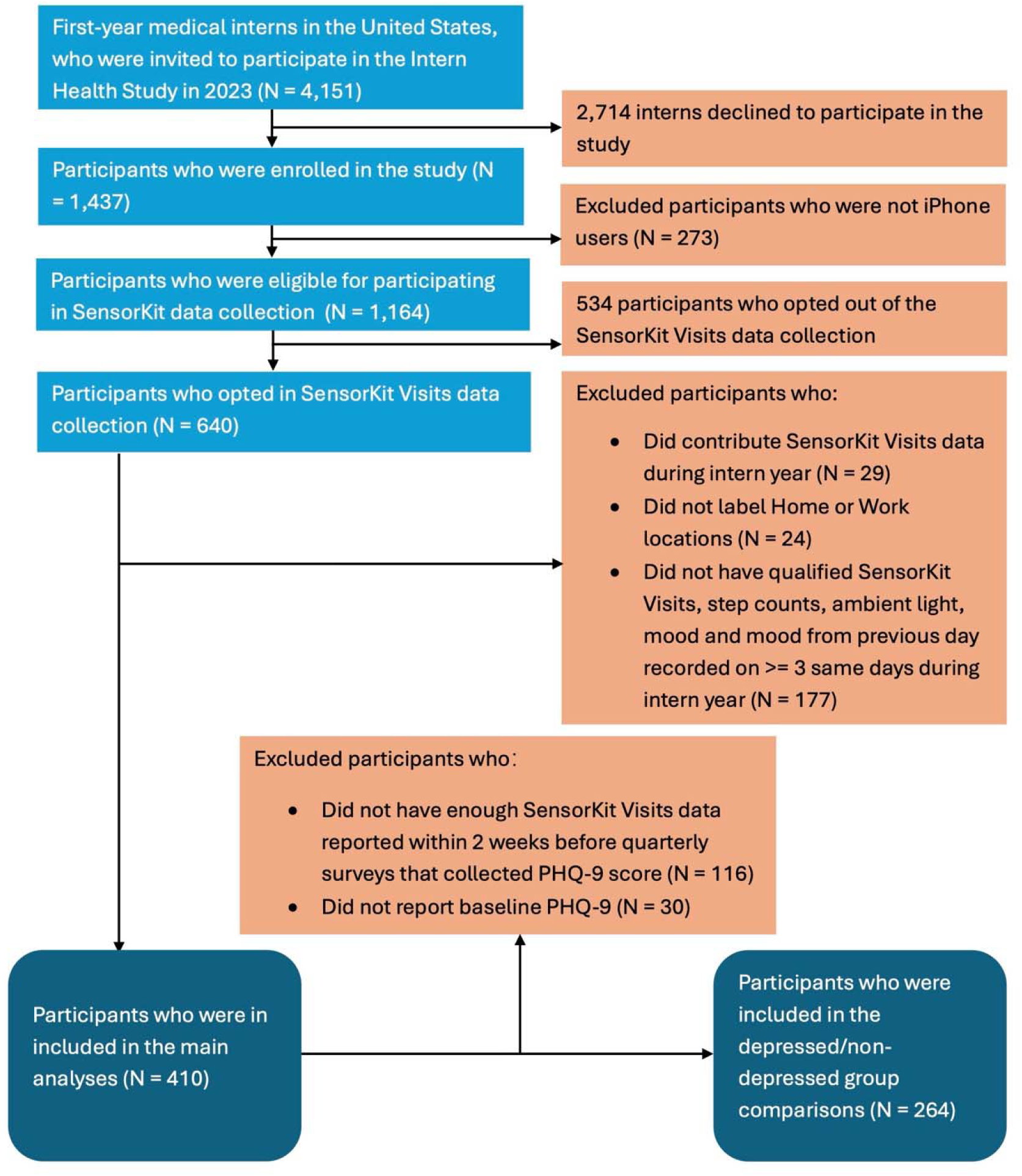
Study Flow Chart.

Across the sample, 18,795 person-days of complete data were collected. On average, subjects contributed 45.8 days of data (SD=42.1, range 3-162, median=29.5). Time spent at third places spanned the full range from 0% to 100%, with an overall mean of 17.9% (SD=26.5%, median=6.3%) and subjects visited 3.3 unique places (SD=1.7, range 1-17, median=3) on average. Mood ratings across all observations averaged 7.0 (SD=1.5, range 1-10, median=7.0). **Figure 2** shows the distributions of the two daily visit metrics and daily mood ratings across the entire sample.

**Figure 2.**
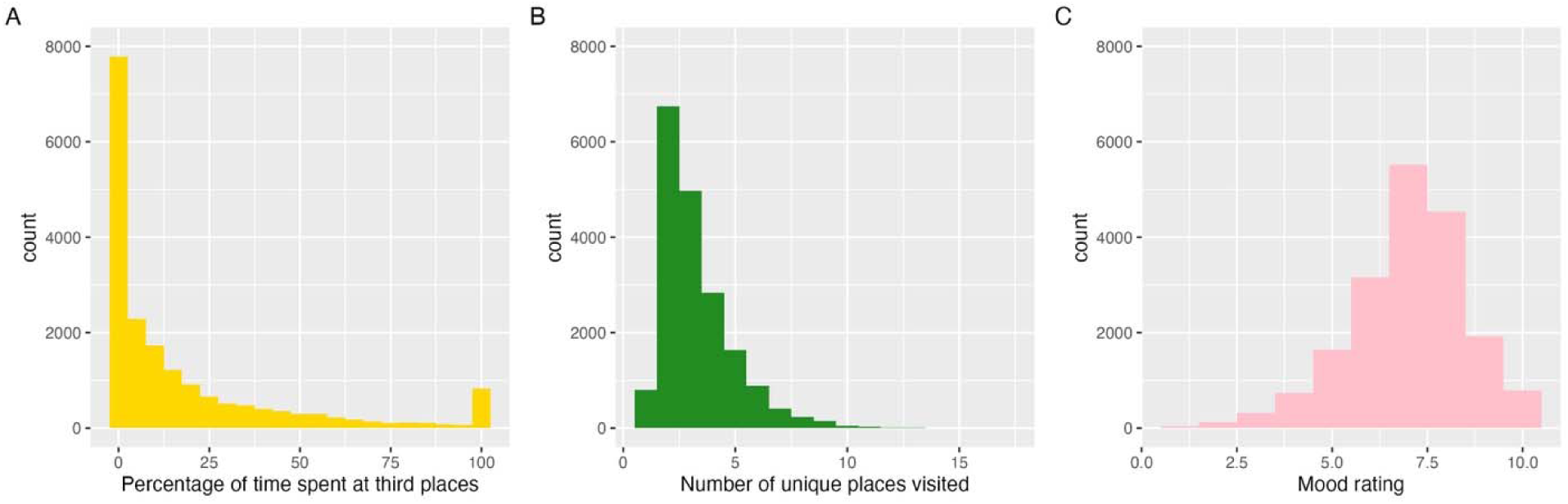
The distributions of (A) percentage of time spent at third places, (B) number of unique places visited per day, and (C) daily mood ratings.

The average individual mean internship PHQ-9 score was 5.8 (SD = 3.9; range: 0.0-22.0; median = 5.0). Among demographics variables, female sex and surgical specialty were significantly associated (p < 0.05) with fewer third places visits and therefore were included as covariates in the analytical models to adjust for potential confounding.

Figure S1 illustrates an example of the raw visits data, represented by categories of visited places and the most visited unique places, from one participant across the entire internship year. **Figure S2** depicts the two derived daily visit metrics (percent time spent in third places, and the number of unique places visited) from the same subject calculated from the raw visits data.

### Associations between daily mood and third places visits

Overall, spending a higher percentage of time in third places and visiting a greater number of unique locations were both associated with better same-day mood (Figure 3). Specifically, a greater percentage of time spent in third places (effect size per 10% increase, same below) was significantly associated with improved mood (between subjects: B = 0.085, p = 0.001; within subjects: B = 0.040, p < 0.001; **Table 1A, Model 1**). These associations remained significant after adjusting for time spent at work (between subjects: B = 0.069, p = 0.007; within subjects: B = 0.022, p < 0.001; **Table 1A, Model 2**). Similarly, visiting a greater number of unique locations was significantly associated with improved mood (between subjects: B = 0.125, p = 0.003; within subjects: B = 0.058, p < 0.001; **Table 1B, Model 1**). These associations also remained significant after adjusting for time spent at work (between subjects: B = 0.105, p = 0.014; within subjects: B = 0.040, p < 0.001; **Table 1B, Model 2**). When both predictors were included in the model, only the within-subject effects remained significant (percentage of time in third places: B = 0.013, p = 0.001; number of unique places visited: B = 0.032, p < 0.001; **Table 1C, Model 2**).

**Figure 3.**
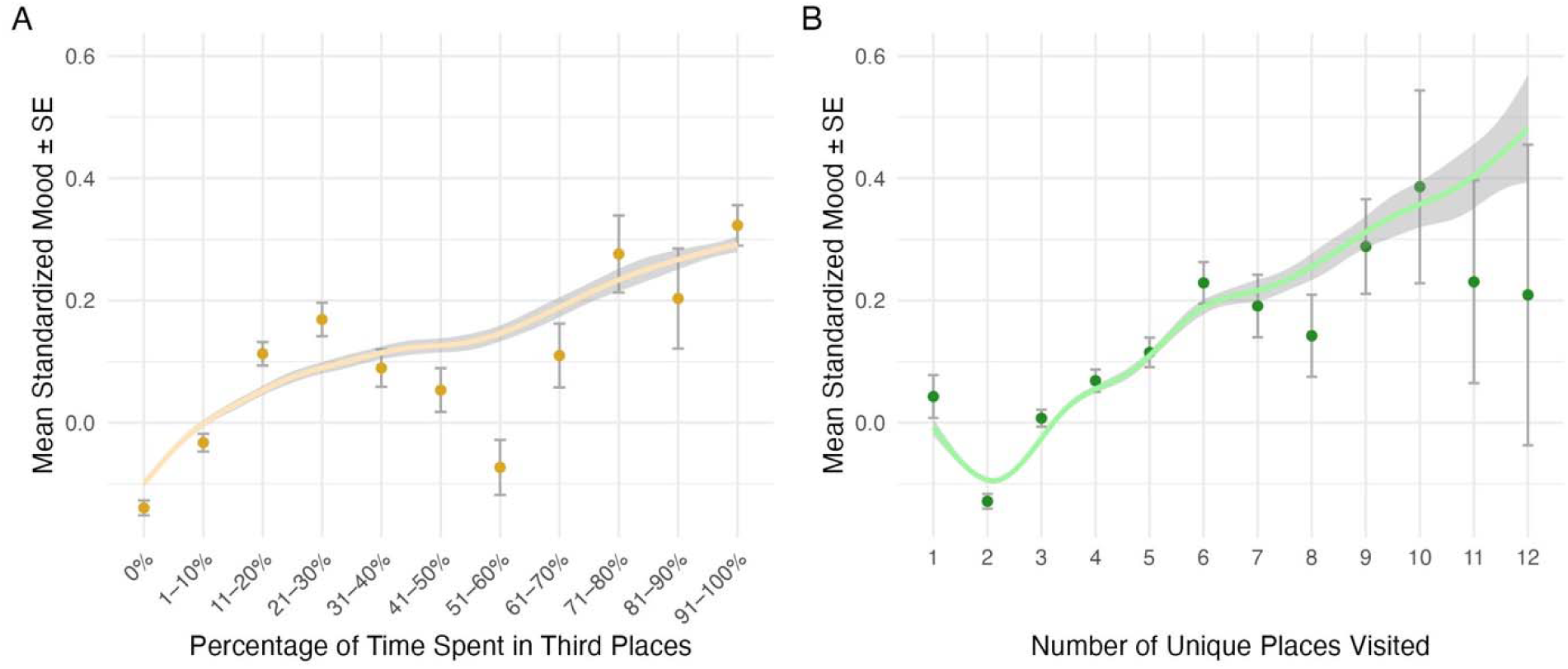
Average daily mood by (A) percentage of time spent in third places and (B) number of unique places visited. Mood was converted to individual z-scores. Percentages of time spent in third places are grouped in 10% increments for clarity. Values for >12 places visited were excluded due to small sample size (<10 per value) and high variance. The GAM-smoothed prediction lines from covariate-adjusted linear mixed models were plotted. Error bars represent standard errors.

**Table 1.**
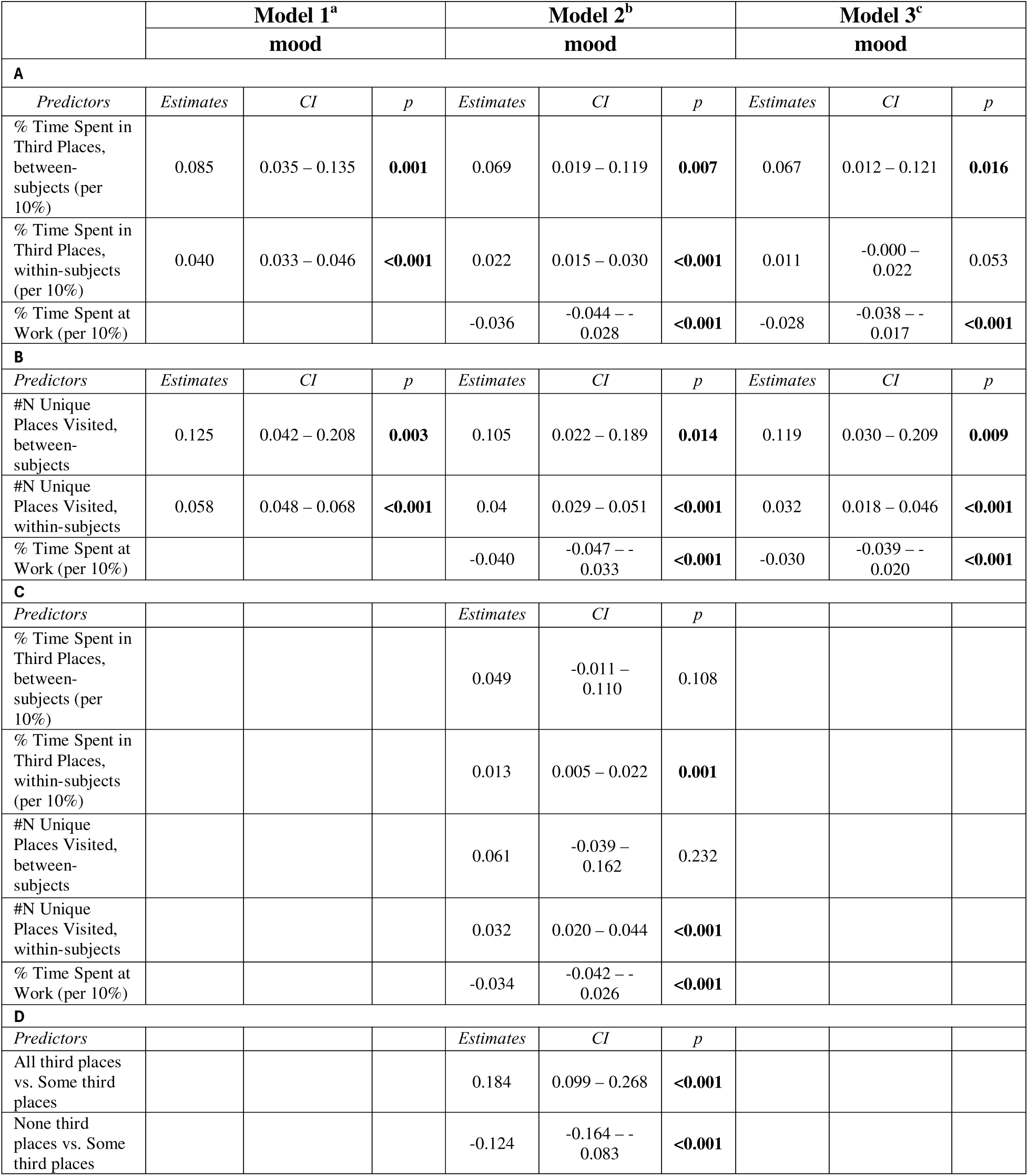
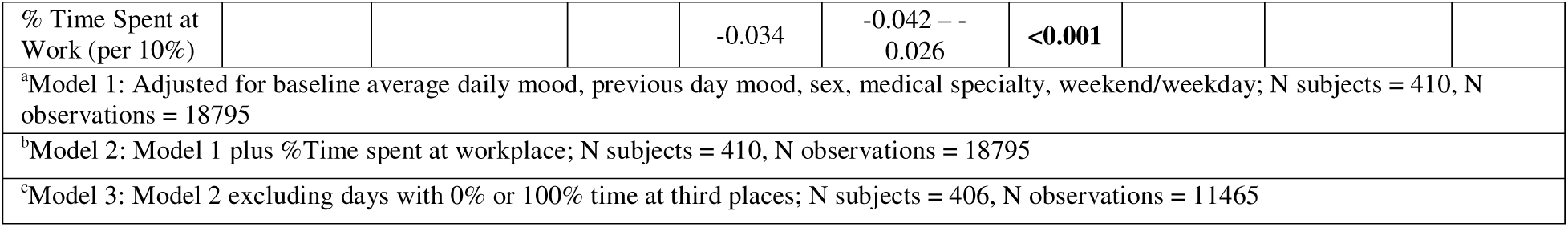
The associations between third places visits and same day mood. The predictors include: A) percentage of time spent in third places; B) number of unique places visited; C) both percentage of time spent in third places and number of unique places visited; and D) categorical variables representing “No third places” (0% of time in third places), “All third places” (100% of time in third places), and “Some third places” (>0% but <100% of time in third places, used as the reference level).

Participants reported significantly lower mood on days when they did not visit any third places (B = – 0.124, p < 0.001) and significantly higher mood on days spent exclusively in third places (e.g., while on vacation; B = 0.184, p < 0.001; **Table 1D, Model 2**), compared to days when part of their time was spent at third places. After excluding days with no third places visits or days spent entirely in third places, the association between the percentage of time spent in third places and mood was significant only at the between-subjects level (B = 0.067, p = 0.016; **Table 1A, Model 3**), while number of unique locations remained significantly associated with mood on both between-subjects (B=0.119, p=0.009) and within-subjects (B=0.032, p<0.001) levels (**Table 1B, Model 3**).

Conversely, within subject, previous day mood was positively associated with both the percentage of time spent in third places (B=0.052, *p*<0.001; B=0.039, *p*<0.001 when excluding days with 0% or 100% time spent in third places) and the number of unique places visited (B=0.032, *p*<0.001) on the following day (**Table 2**).

**Table 2.** The associations between previous day mood and third places visits, including percent of time spent in third places (left and middle columns) and number of unique places visited (right column). Both outcome variables were rank-based inverse normal transformed.

|  | % Time Spent in Third Places<sup>a</sup> |  |  | % Time Spent in Third Places<sup>b</sup> |  |  | #N Unique Places Visited<sup>a</sup> |  |  |
| --- | --- | --- | --- | --- | --- | --- | --- | --- | --- |
| *Predictors* | *Estimates* | *CI* | *p* | *Estimates* | *CI* | *p* | *Estimates* | *CI* | *p* |
| Previous Day Mood, between-subjects | 0.014 | -0.020 – 0.048 | 0.427 | -0.000 | -0.038 – 0.038 | 0.996 | 0.031 | -0.002 – 0.064 | 0.07 |
| Previous Day Mood, within-subjects | 0.052 | 0.041 – 0.062 | **<0.001** | 0.039 | 0.024 – 0.054 | **<0.001** | 0.032 | 0.021 – 0.043 | **<0.001** |
| <sup>a</sup> Adjusted for sex, medical specialty, weekend/weekday; N subjects = 410, N observations = 18795 |  |  |  |  |  |  |  |  |  |
| <sup>b</sup> Same covariates as a, and excluding days with 0% or 100% time at third places; N subjects = 406, N observations = 11465 |  |  |  |  |  |  |  |  |  |

### Mediation effects of physical activity and outdoor light exposure on the relationship between third places visits and mood

Daily step count (per 1000 steps) was significantly associated with daily mood (B=0.016, p<0.001), percentage of time spent at third places (B=0.189, p<0.001), and number of unique places (B=0.762, p<0.001). Step count explained 10.7% (95%CI: 6.6%-17.3%) of the association between percentage of time spent at third places and mood, and 18.2% (95%CI: 10.7%-28.6%) of the association between number of unique places and mood (Figure 4C and D).

**Figure 4.**
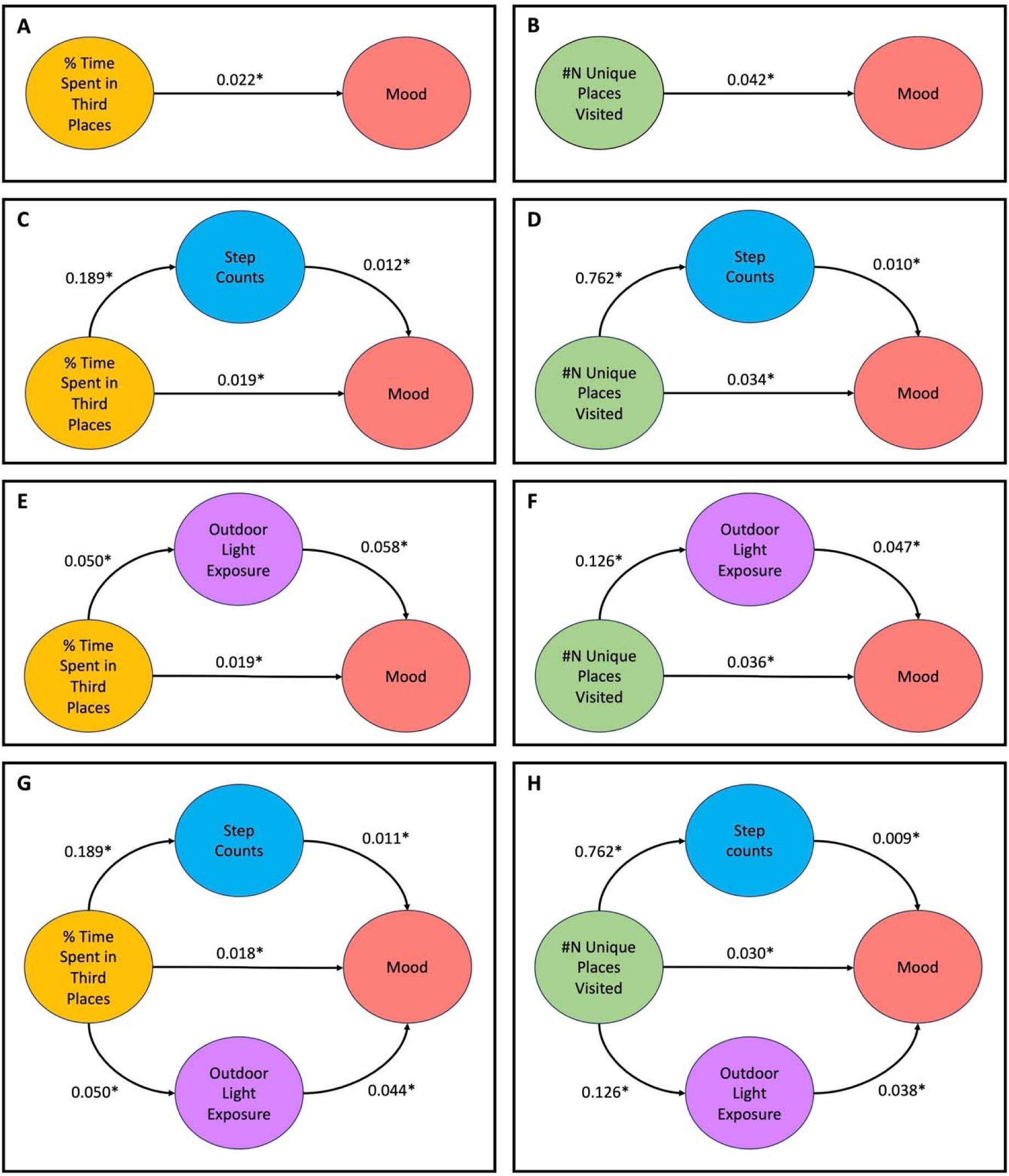
Mediations of the associations between third places visits and same day mood by same day step counts and outdoor light exposure. A–B, The associations of third places visits (percentage of time spent in third places, and number of unique places visited) and same day mood; C–D, The associations mediated by same day step counts; E–F, The associations mediated by same day outdoor light exposure; G–H, the associations mediated jointly by same day step counts and outdoor light exposure. The effect sizes reflect the estimated change associated with a 10% increase for percentage time spent in third places and outdoor light exposure (represented by the percentage of time with light exposure illuminance levels >= 1000 lux). * all P□<□1□×□10^−3^.

Similarly, daily outdoor light exposure, operationalized as daily percentage of time with illuminance >= 1000 lux (effect size per 10% increase, same below), was significantly associated with daily mood (B=0.079, p<0.001), percentage of time spent at third places (B=0.050, p<0.001), and number of unique places (B=0.126, p<0.001). Outdoor light exposure explained 13.2% (95%CI: 7.1%-22.7%) of the association between percentage of time spent at third places and mood, and 14.2% (95%CI: 6.7%-24.0%) for number of unique places and mood (Figure 4E and F).

Jointly, daily step count and daily outdoor light exposure explained 19.2% (95%CI: 12.0%-31.5%) of the association between percentage of time spent at third places and mood, and 27.6% (95%CI: 17.2%-43.0%) for number of unique places and mood (Figure 4G and H).

### Comparison of third places visits between subjects who met criteria for depression and those who did not

To assess between-subject associations with third places visits, we compared the percentage of time spent in third places and the number of unique places visited between the subjects who met criteria for depression (whose average internship PHQ-9 scores >= 10, N = 50) and those who did not (N = 214). On average, depressed subjects spent significantly less time in third places (12.3% vs. 21.2%, t=-3.7, *p*<0.001) and visited significantly fewer unique places (2.9 vs. 3.4, t = –3.8, *p*<0.001) than non-depressed subjects (Figure 5). After adjusting for baseline PHQ-9 scores, sex, and medical specialty, the estimated marginal means between depressed and non-depressed groups remained significant (percentage of time spent at third places: 9.7% vs. 19.8%, t=-3.1, *p*=0.003; number of unique places visited: 2.8 vs. 3.4, t = – 2.7, *p*=0.007).

**Figure 5.**
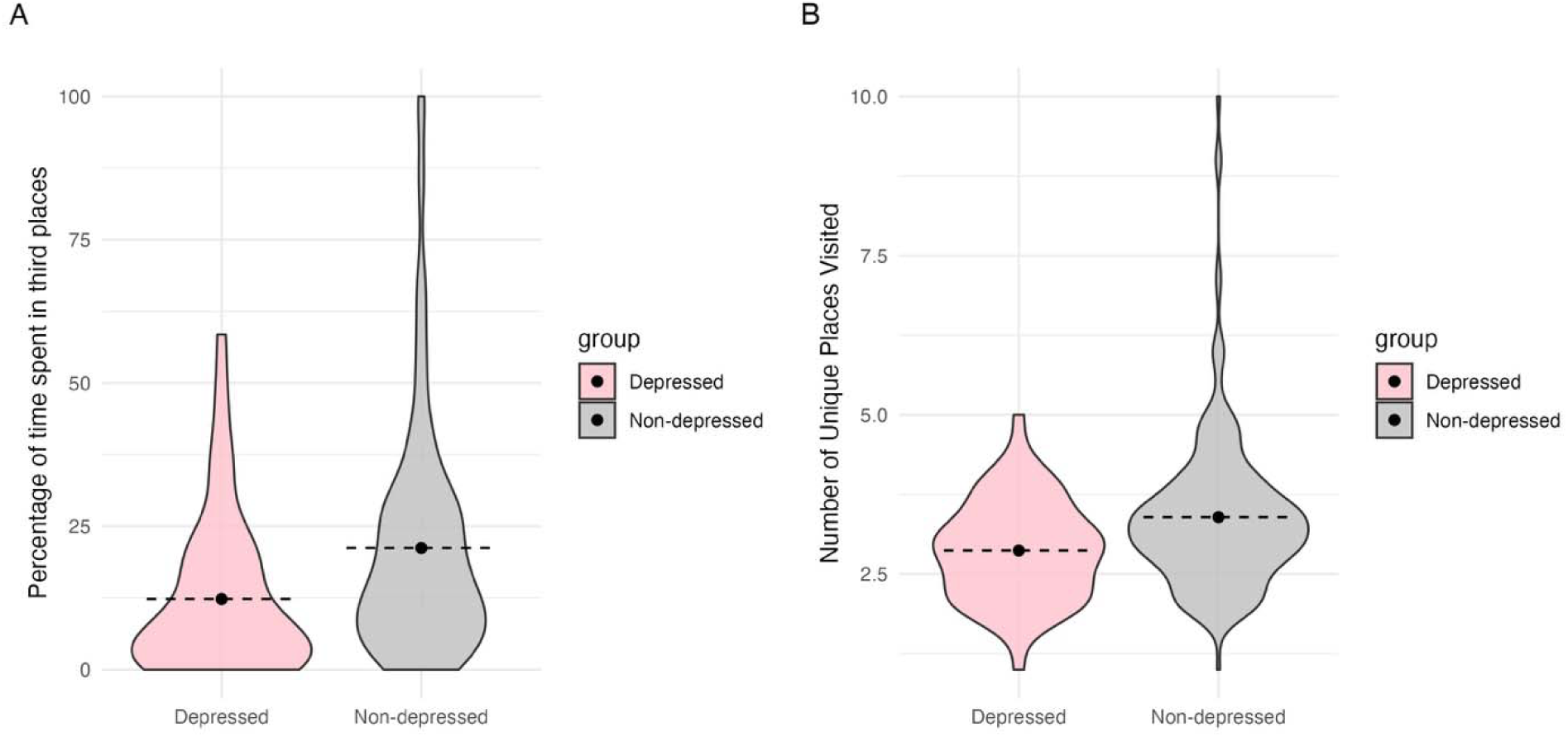
Comparisons of third places visits between depressed (N=50) and non-depressed (N=214) subjects.

### Sensitivity analysis excluding records from midnight to 6am

We conducted a sensitivity analysis excluding records from midnight to 6 a.m. (18,725 observations from 408 subjects). The results were similar to the main analysis (**Tables S2, S3**; **Figure S3**).

## Discussion

Utilizing longitudinal, passively collected SensorKit visits data from a national cohort of first-year medical residents across a full year, we found that a higher percentage of time spent at third places and a greater number of unique locations visited were consistently linked to better subsequent mood, both between and within subjects. Furthermore, our findings support the presence of the reverse relationship between mood and third places visits: experiencing better mood on one day was associated with increased time spent at third places and a greater number of unique places visited on the following day. This dynamic underscores the possibility that third places visits and positive affect may reinforce one another over time. Additionally, we found that subjects meeting PHQ-9 criteria for depression spent significantly less time in third places and visited significantly fewer unique locations than their non-depressed peers, providing further support for the link between limited third places visits and depression.

Work hours are a strong predictor of depressive symptoms among interns^41^ and also limit the time available for third places visits. Thus it could seem plausible that the positive effect of third places visits on mood was simply the result of spending less time at work. However, even after adjusting for time spent at the workplace, the association between both the percentage of time spent at third places and the number of unique places visited and mood remained significant. This finding suggests that visiting third places is linked to better mood independent of work hours and that the benefits of third places go beyond simply reducing time spent at work.

Several mechanisms may mediate the association between third places visits and mood. For instance, third places can provide a sense of community and social connection^4,6,44,45^, which supports psychological health^46^. Visiting third places may also increase physical activity^20,25^ which has been shown to benefit the mental health of populations under stress^22^. Additional factors such as increased outdoor light exposure, engagement in leisure activities, and opportunities for restorative experiences may also contribute. Based on the data availability of the Intern Health Study, we tested the mediation effects of two potential mechanisms, physical activity and outdoor light exposure, and found that both significantly mediated the associations between third places visits and mood. Notably, these two factors jointly explained 19.2%-27.6% of the associations, suggesting that exercise and light exposure are meaningful mediating factors, but that other mechanisms are likely also involved. Further research should investigate additional potential mediators as more data become available.

Taken together, these findings provide valuable real-world, empirical support to the theoretical foundation linking third places visits to mood and depression. Unlike prior studies that often relied on self-reported data, our approach utilized objective, quantitative and precise location measures collected passively and unobtrusively by SensorKit gathered from a large national cohort. The larger sample and objective, comprehensive assessment of location offers a deeper and more comprehensive understanding of how engagement with third places relates to mental health in everyday life.

Further, the longitudinal nature of our study, following subjects across one full year, allowed us to examine within-subject associations over time, overcoming the limitations of cross-sectional studies. By tracking individuals’ behaviors and mood fluctuations throughout the year, we were able to identify the dynamic links between third places visits and mood at the individual level, rather than solely assessing between-person differences. We found that changes in third places engagement leads to changes in mood, and vice versa, within the same person, strengthening the understanding of directional effects and reducing confounding from individual differences. These findings can also inform individual interventions and public health strategies.

Finally, the operational definition of “third place” in this study encompassed all non-work, non-home settings, addressing a key limitation of earlier studies that focused on a restricted set of locations. By capturing a broad spectrum of third places experiences across an extended period, our findings are more generalizable and valid.

In addition to these strengths, our study has several limitations. First, while the analyses adjusted for key demographic, baseline and time-varying variables and were longitudinal in nature, the observational nature of our study precludes causal inference. Second, missing or inaccurate data due to periods when phones were turned off or left behind could potentially introduce bias. Third, our sample was limited to iPhone users who consented to passive location data collection, which may introduce selection bias and affect the generalizability of our findings. Finally, although the stressors of medical internship provide a useful framework to examine potential protective factors for depression, findings from this unique population of training physicians need to be examined in other populations to validate its generalizability.

In conclusion, our study employs innovative measurement approaches and a longitudinal study design to advance our understanding of the relationship between third places and well-being. We provide robust evidence that everyday engagement with places outside home and work is associated with better mood and less depressive symptoms and sheds light on some of the mechanisms that underlie the connection. This work has the potential to inform evidence-based interventions utilizing third places to promote psychological well-being.

## Data Availability

SensorKit data cannot be shared publicly because of Apple's data sharing restrictions.
De-identified general study enrollment and survey data are available via the ICPSR repository (https://www.openicpsr.org/openicpsr/project/129225/version/V1/view).
De-identified wearable data is available through requests to the Intern Health Study. Reasonable requests will be fulfilled contingent on completion of a data use agreement with the University of Michigan.

