## Supplementary figures and images for "Third places visits and well-being: insights from longitudinal passive sensing data"

### Figure S1

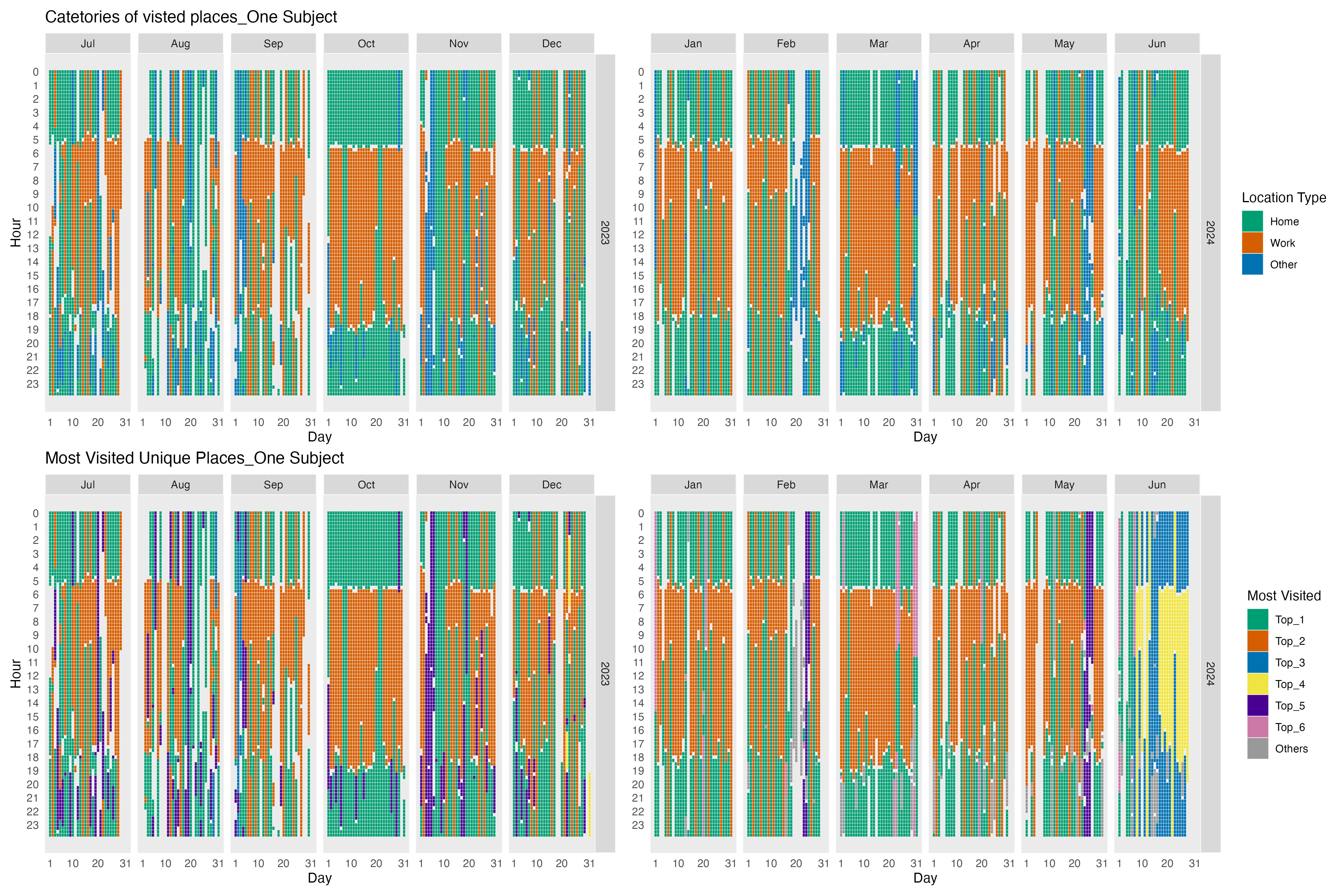

### Figure S2

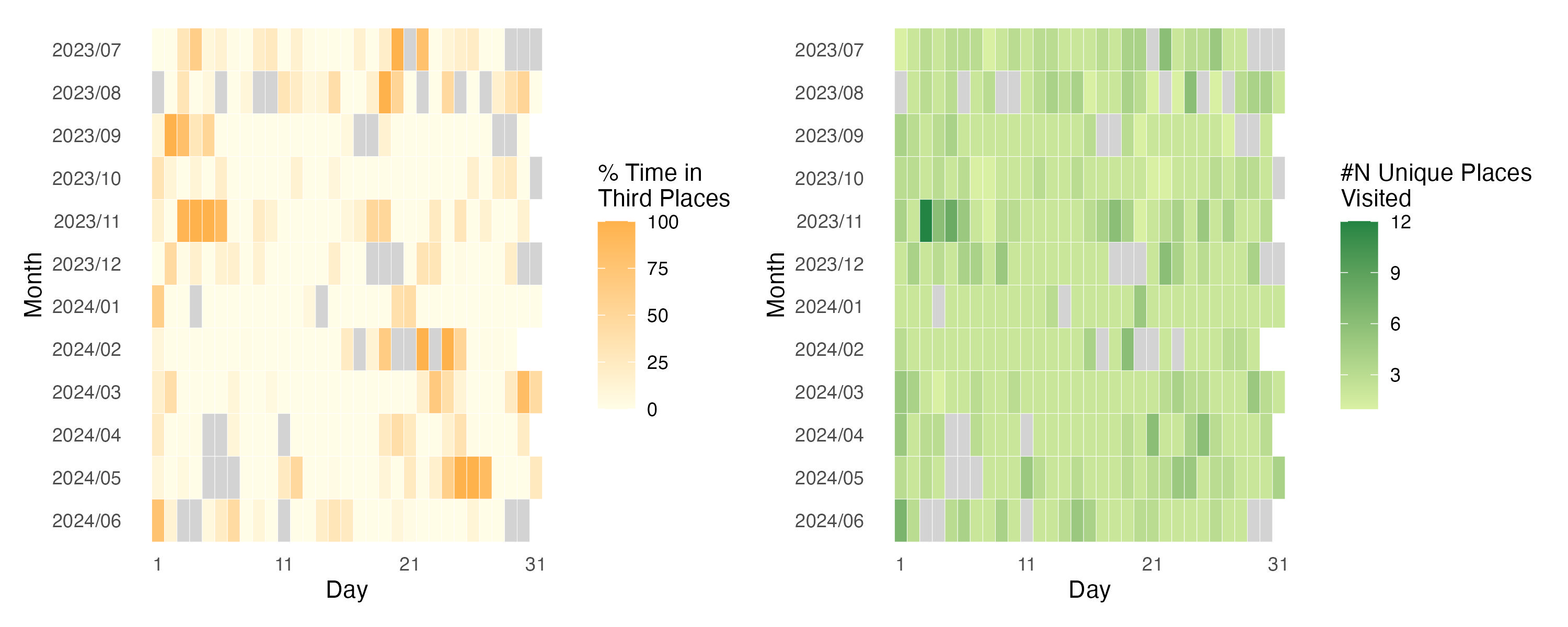

### Figure S3

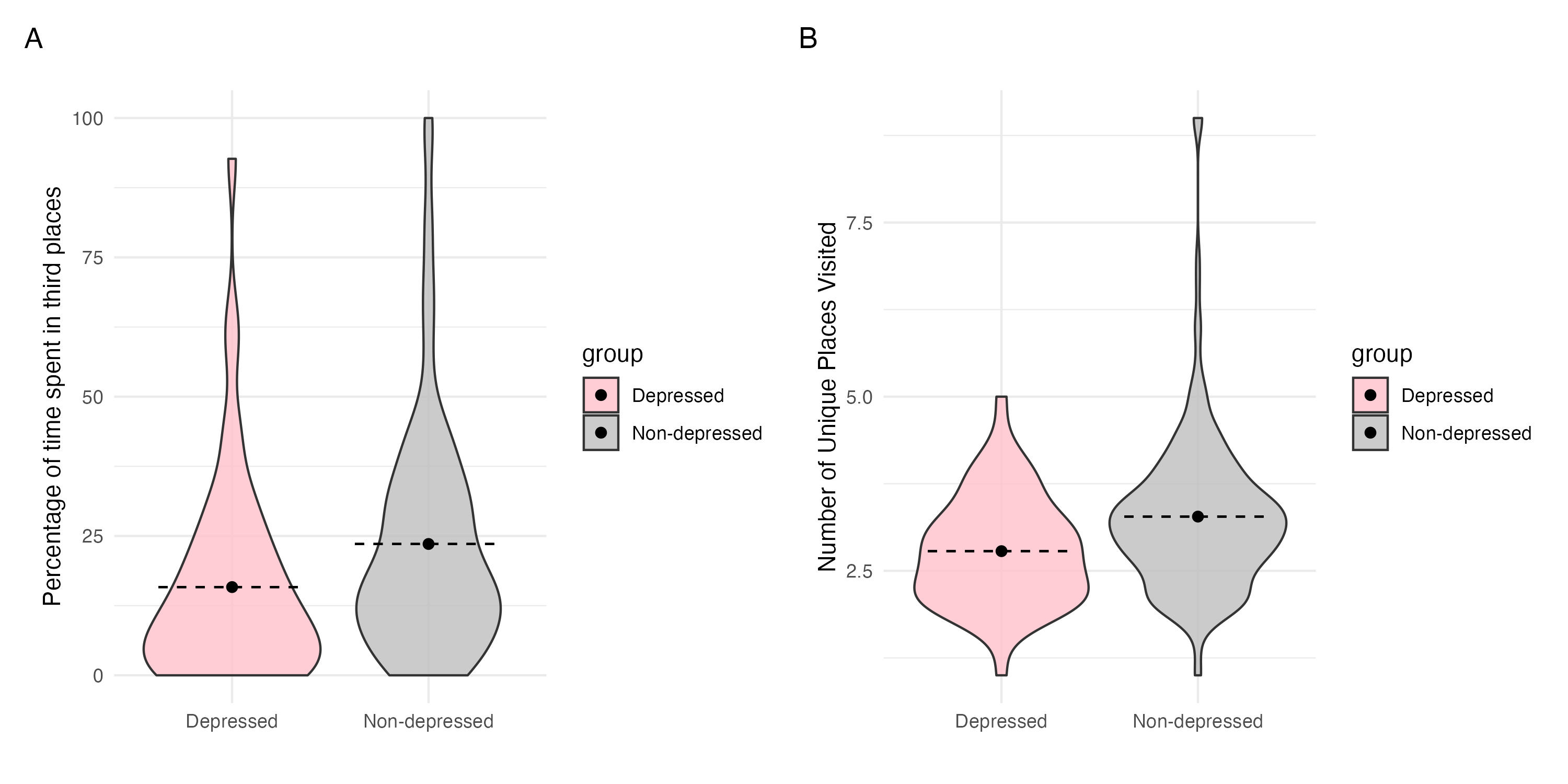
