## Supplementary material for "Third places visits and well-being: insights from longitudinal passive sensing data": Table S1

**Table S1. SensorKit Visits data collection**

|  | **DATA COLLECTED** | **DATA NOT COLLECTED** |
| --- | --- | --- |
| **Visits** | - Location category (e.g. home, work, school) if known - Anonymized identifier of frequently visited locations - The distance of an anonymized location from home. - Arrival and departure times that are expanded to 15-minute increments | - GPS or specific location information - Names of the places visited - Addresses of the places, including home or work |
